# Large-Scale Multi-Omics Enhance Risk Prediction for Type 2 Diabetes

**DOI:** 10.64898/2026.02.19.26346636

**Authors:** Ruijie Xie, Christian Herder, Ben Schöttker

## Abstract

**Introduction:** Polygenic risk scores (PRS), metabolomics, and proteomics have each shown promise in improving type 2 diabetes risk prediction, but their combined utility beyond established clinical models remains unclear. We aimed to evaluate whether integrating multi-omics biomarkers enhances 10-year type 2 diabetes risk prediction beyond single-omics extensions and the clinical Cambridge Diabetes Risk Score (CDRS), which includes HbA_1c_ measurements.

**Methods:** We analysed data from 23,325 UK Biobank participants without diagnosed diabetes at baseline. Data for a PRS for type 2 diabetes, 11 metabolites, and 15 proteins were added to the CDRS to develop multi-omics prediction models. Model performance was evaluated using Harrell’s C-index and the net reclassification index (NRI).

**Results:** During 10 years of follow-up, 719 participants developed incident type 2 diabetes. Among individual omics layers, proteomics contributed the greatest improvement in predictive performance, increasing the C-index from 0.857 (clinical CDRS) to 0.880 (ΔC-index; +0.023; *P* < 0.001), with an NRI of 30.0%. The full multi-omics model, further significantly increased the C-index compared to a model combining the clinical CDRS with proteomics data (C-index, 0.886; ΔC-index; +0.006; *P* < 0.033).

**Conclusion:** Integrating proteomics, metabolomics, and a diabetes-PRS into a clinical model substantially improves type 2 diabetes risk prediction beyond single-omics extensions. However, the C-index difference between the proteomics extended and full multi-omics extended models is small, and the clinical models extended with proteomics data would be easier to translate into routine care because it needs only the measurement of 15 proteins.

## 1. Introduction

Type 2 diabetes is a major global health concern, with its rising prevalence leading to increased morbidity, premature mortality, and substantial economic burden [1–3]. Early identification of individuals at high risk is essential for implementing targeted preventive strategies to delay or prevent disease onset [4, 5]. However, existing prediction models often lack specificity and fail to capture the complex interplay of biological and environmental factors contributing to type 2 diabetes development, thereby limiting their clinical utility [6, 7].

To address these limitations, genomics, metabolomics, and proteomics have been increasingly explored as complementary tools for risk prediction [8–10]. Genome-wide association studies (GWAS) have identified numerous genetic variants associated with type 2 diabetes, while metabolomics and proteomics have uncovered key pathways involved in insulin resistance, inflammation, and metabolic dysfunction [11–13]. However, polygenic risk scores (PRS) have generally shown limited added value over traditional clinical models, and although metabolomics and proteomics have each demonstrated promising results, their combined predictive utility remains insufficiently explored [14–17]. To date, only one study has systematically assessed the integration of genomics, metabolomics, and proteomics for type 2 diabetes prediction in a multi-omics approach [18]. In the EPIC-Norfolk study (N = 1,105), Zanini et al. reported that integrating the top 10 features from genomics, metabolomics, and proteomics significantly improved 10-year type 2 diabetes prediction over a clinical model alone (C-index: 0.82 vs. 0.87) [18]. However, large-scale, population-based studies are still lacking to assess the comparative and joint predictive value of different omics layers, and to identify the optimal omics combination for improving type 2 diabetes risk prediction.

In previous research using the UK Biobank (UKB), our group developed proteomics-based and metabolomics-based type 2 diabetes risk models that significantly improved the predictive performance of the clinical Cambridge Diabetes Risk Score (CDRS) [19, 20]. The CDRS is a sex-independent model that incorporates age, sex, body mass index (BMI), family history of diabetes, smoking status, and use of antihypertensive and steroid medications, with optional inclusion of glycated hemoglobin (HbA_1c_) [18, 21]. Separately, the UKB has also released a genome-wide polygenic risk score for type 2 diabetes (T2D-PRS) ¹.

We aim to build upon this previous research on the single omics layers and combine the selected features from proteomics [19], metabolomics [20], and genomics [22] with the clinical CDRS model [18, 21] to determine the optimal combination of omics layers for 10-year type 2 diabetes risk prediction.

## 2. Methods

### 2.1 Study population

The UKB is a large prospective cohort comprising 502,414 participants aged 37 to 73 years, recruited from 13 March 2006 to 1 October 2010 across 22 assessment centers in England, Scotland, and Wales [23]. The cohort includes extensive phenotypic and genetic data, encompassing blood and urine biomarkers, whole-body imaging, lifestyle factors, anthropometric measurements, and genomic sequencing. UK Biobank received ethical approval from the North West Multi-centre Research Ethics Committee (REC reference 11/NW/0382). This study was conducted under UK Biobank application number [101633].

The starting point for the current analysis was a subsample of 54,219 participants with available proteomics measurements. We excluded participants with more than 50% missing protein measurements (n=1,871), and those who lacked metabolomics data (n=22,417) or PRS data (n=193), resulting in 29,738 participants with complete multi-omics data. Participants with diagnosed type 2 diabetes (n=1,733), with potential undiagnosed type 2 diabetes, defined as HbA_1c_ ≥ 6.5% according to the American Diabetes Association (ADA) criteria (n=1,552) [24], or missing diabetes status (n=118) were further excluded. Finally we excluded participants who were not randomly selected for the proteomics measurements (n=3,010). Ultimately, 23,325 participants were included in the final analysis (**Supplemental Figure S1**).

### 2.2 Reference model: The clinical CDRS

The CDRS is a well-established tool for predicting future risk of type 2 diabetes. It incorporates age, sex, BMI, family history of diabetes, smoking status, and the use of antihypertensive and steroid medications [21]. In this study, we used the clinical version of the CDRS that additionally includes HbA_1c_ levels [18]. In the UK Biobank, age, sex, family history of diabetes, and smoking status were collected through standardized questionnaires. Information on prescribed medications was obtained through self-reported data, verbal interviews, and linkage with primary care records. Anthropometric measurements, including weight and height, were performed by trained staff at assessment centers. HbA_1c_ levels were measured from whole-blood samples using high-performance liquid chromatography on the Variant II platform (Bio-Rad Laboratories) [25].

### 2.3 Proteomics data

Proteomic profiling was conducted on EDTA-plasma samples collected at baseline, mostly in a non-fasting state. The assay protocols, including sample handling and selection procedures, have been described previously [26]. Briefly, Olink Proteomics applies the proximity extension assay (PEA) technology, which uses pairs of antibodies linked to complementary oligonucleotides to detect target proteins [27–29]. A total of 2,923 unique proteins were measured using the Olink Explore 3072 platform (Olink Proteomics, Uppsala, Sweden). As outlined in our prior study [19], proteins with more than 20% missing values or over 25% of values below the detection limit were excluded, resulting in a final panel of 2,085 proteins for biomarker selection. In our previous UKB data analysis [19], we applied least absolute shrinkage and selection operator (LASSO) regression with bootstrap resampling to identify a parsimonious set of 15 proteins with high predictive value for type 2 diabetes (**Supplemental Table S1**). Incorporating these proteins into the clinical CDRS model significantly improved risk discrimination, with the C-index increasing from 0.831 to 0.860 (*P* < 0.001). In the present analysis, we evaluated only these 15 preselected proteins.

### 2.4 Metabolomic data

Plasma metabolomic profiling was conducted on EDTA samples, mostly collected in a non-fasting state, using a high-throughput nuclear magnetic resonance (NMR) platform developed by Nightingale Health [30]. This platform quantified 249 circulating metabolites, encompassing a broad spectrum of molecular classes such as lipids, fatty acids, amino acids, ketone bodies, and other low-molecular-weight compounds. Among these, 168 metabolites were measured in absolute concentrations, including 61 composite biomarkers derived from 107 directly quantified metabolites. The remaining 81 metabolites were expressed as concentration ratios. In our previous UKB data analysis [20], we applied LASSO regression with bootstrap resampling to identify a parsimonious set of 11 metabolites with strong predictive value (**Supplemental Table S1**). Incorporation of these metabolites into the clinical CDRS model significantly improved risk discrimination, with the C-index increasing from 0.815 to 0.834 (*P* < 0.001). In the present analysis, only these 11 preselected metabolites were evaluated.

### 2.5 Polygenic risk score for type 2 diabetes

The PRS for type 2 diabetes was provided by the UK Biobank PRS Release [22]. This score was derived using a Bayesian modelling approach, trained on meta-analysed summary statistics from 5 external GWAS datasets, and subsequently optimized using internal UK Biobank data. In internal benchmarking analyses, the UKB T2D-PRS demonstrated strong predictive performance and was shown to be comparable or superior to previously published PRSs (e.g., Mahajan et al. and Khera et al.) [22, 31, 32].

### 2.6 Type 2 diabetes ascertainment

Participants were censored at the earliest of the following events: diagnosis of type 2 diabetes, death, loss to follow-up, or completion of the 10-year follow-up period. Incident type 2 diabetes was identified using three complementary data sources in the UKB [33]. First, self-reported diagnoses and the use of glucose-lowering medications were collected during follow-up verbal interviews and questionnaires. Second, primary care records, hospital admission data, and death registry data were searched for relevant medical codes indicative of type 2 diabetes. Third, prescription records from primary care were reviewed, and participants with prescriptions for glucose-lowering medications (Anatomical Therapeutic Chemical [ATC] classification code group A10) were classified as incident cases.

### 2.7 Statistical analyses

#### 2.7.1 General remarks

All statistical analyses were conducted in R software (version 4.4.0, R Foundation for Statistical Computing, Vienna, Austria). Statistical significance was set at *P* < 0.05 (two-sided). Missing values of variables of the clinical CDRS (variable with the highest proportion of missing values was family history of diabetes with 2.3%) and multi-omics (mostly complete with a few proteins with up to 20% of missing values) were single imputed using the chained equations method with random forest algorithms implemented in the R package *missForest* (version 1.5) [34].

#### 2.7.2 Test of Model Performance

The study design is illustrated in **Graphical abstract**. Panels of 15 proteins, 11 metabolites, and the UK Biobank’s T2D-PRS, derived in previously published selection procedures [19, 20, 22], were combined with the variables of the clinical CDRS to develop a 10-year type 2 diabetes risk prediction model [21]. The UKB cohort was randomly split into a training set (70%) and a test set (30%) for model development and validation.

Model discrimination was evaluated using Harrell’s C-index. Differences in C-index between models were assessed using the method by Kang et al. for comparing correlated C-indices in survival analysis, implemented in the R package *compareC* (version 1.3.2) [35]. The incremental C-statistic of each selected biomarker was also estimated. In a first step, we assessed the predictive performance of each individual omics layer when added to the clinical CDRS model. In a second step, the combination of the best performing omics layer and the clinical CDRS was selected as the reference model and we tested whether adding more omics layers improved its predictive ability.

Risk reclassification was evaluated using the continuous net reclassification index (NRI) and the integrated discrimination index (IDI) [36, 37]. Model calibration was assessed by plotting observed 10-year type 2 diabetes incidence against predicted risks across deciles of absolute risk.

We additionally conducted subgroup analyses for subjects free of prediabetes (HbA_1c_ < 5.7% [<39 mmol/mol]) and those with prediabetes (HbA_1c_ 5.7–6.4% [39–47 mmol/mol]), based on the HbA_1c_ cut-offs defined by the ADA criteria [24].

#### 2.7.3 Associations of selected biomarkers with incident type 2 diabetes

To report hazard ratios (HRs) and 95% confidence intervals (CIs) for the associations of the selected biomarkers (per one standard deviation [SD] increment) with 10-year incident type 2 diabetes, each biomarker was entered separately into Cox proportional hazards models in the validation set (R package *survival* (version 3.5-5)). All models were adjusted for the variables of the clinical CDRS.

#### 2.7.4 Correlation matrix

To assess the independence of the selected multi-omics biomarkers, Spearman correlation coefficients were calculated in the test set.

## 3. Results

### 3.1 Baseline characteristics and incident type 2 diabetes cases

**Table 1** presents the baseline characteristics of 23,325 participants included in the study, of whom 719 developed type 2 diabetes during the 10-year follow-up period. The mean age of the total cohort was 56.4 ± 8.1 years, with 44.5% being male. All features of the clinical CDRS model statistically significantly differed between incident T2D cases and controls. Compared to participants who did not develop type 2 diabetes in the next 10 years, those who did were more likely to be male (56.9% vs. 44.1%), had higher mean age (59.0 vs. 56.3 years), HbA_1c_ levels (5.8% vs. 5.3%), and BMI (30.9 vs. 27.0 kg/m²). A higher proportion of incident T2D cases also had a family history of diabetes (29.2% vs. 17.4%), were smokers or former smokers (54.4% vs 44.1%) and used antihypertensive (15.9% vs. 9.5%) or steroid medications (2.4% vs. 1.2%) at baseline.

**Table 1.**
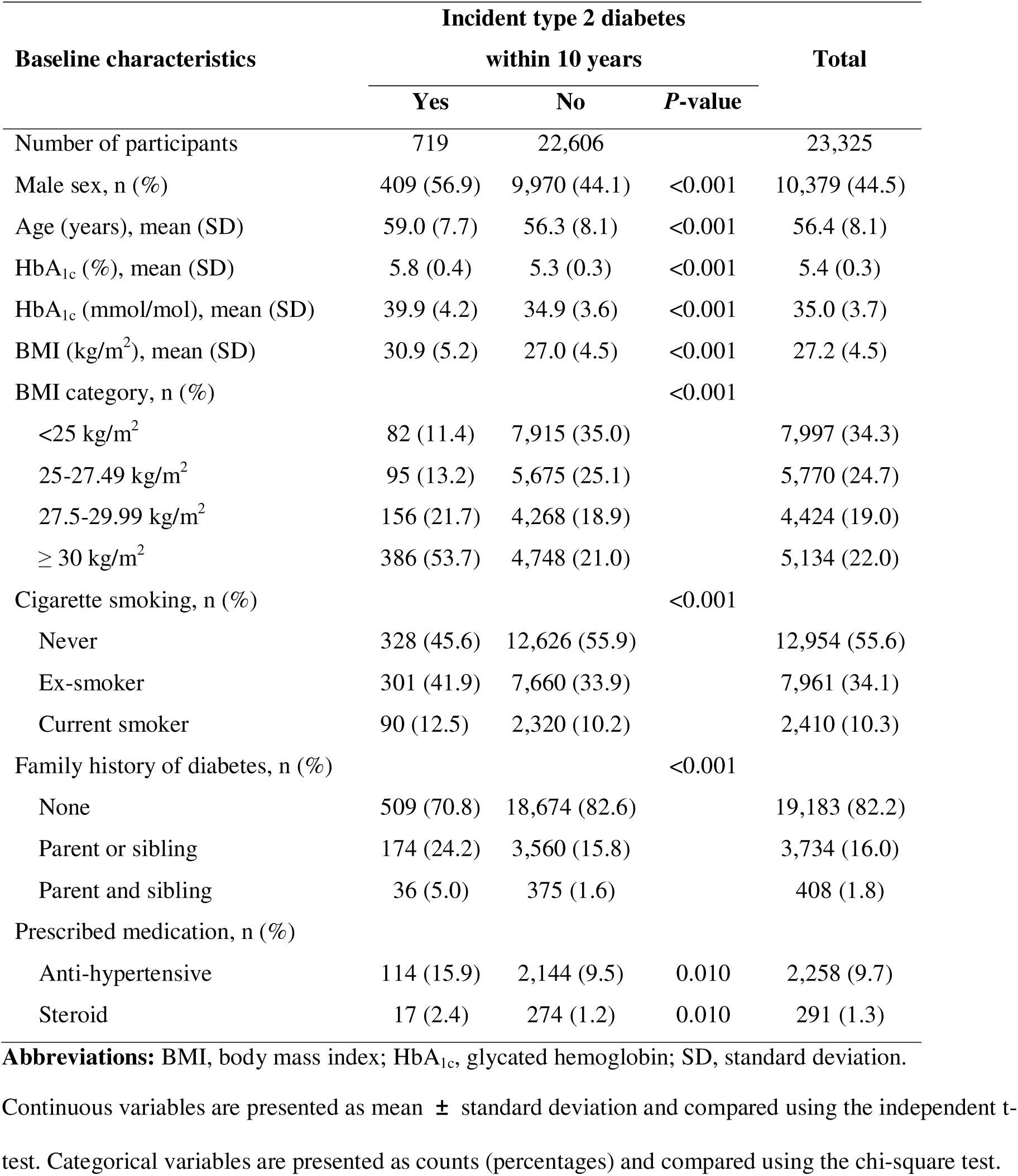
Baseline characteristics of selected participants from the UK Biobank.

| Baseline characteristics | Incident type 2 diabetes |  |  | Total |
| --- | --- | --- | --- | --- |
|  | within 10 years |  | *P*-value |  |
|  | Yes | No |  |  |
| Number of participants | 719 | 22,606 |  | 23,325 |
| Male sex, n (%) | 409 (56.9) | 9,970 (44.1) | <0.001 | 10,379 (44.5) |
| Age (years), mean (SD) | 59.0 (7.7) | 56.3 (8.1) | <0.001 | 56.4 (8.1) |
| HbA<sub>1c</sub> (%), mean (SD) | 5.8 (0.4) | 5.3 (0.3) | <0.001 | 5.4 (0.3) |
| HbA<sub>1c</sub> (mmol/mol), mean (SD) | 39.9 (4.2) | 34.9 (3.6) | <0.001 | 35.0 (3.7) |
| BMI (kg/m<sup>2</sup>), mean (SD) | 30.9 (5.2) | 27.0 (4.5) | <0.001 | 27.2 (4.5) |
| BMI category, n (%) |  |  | <0.001 |  |
| <25 kg/m<sup>2</sup> | 82 (11.4) | 7,915 (35.0) |  | 7,997 (34.3) |
| 25-27.49 kg/m<sup>2</sup> | 95 (13.2) | 5,675 (25.1) |  | 5,770 (24.7) |
| 27.5-29.99 kg/m<sup>2</sup> | 156 (21.7) | 4,268 (18.9) |  | 4,424 (19.0) |
| ≥ 30 kg/m<sup>2</sup> | 386 (53.7) | 4,748 (21.0) |  | 5,134 (22.0) |
| Cigarette smoking, n (%) |  |  | <0.001 |  |
| Never | 328 (45.6) | 12,626 (55.9) |  | 12,954 (55.6) |
| Ex-smoker | 301 (41.9) | 7,660 (33.9) |  | 7,961 (34.1) |
| Current smoker | 90 (12.5) | 2,320 (10.2) |  | 2,410 (10.3) |
| Family history of diabetes, n (%) |  |  | <0.001 |  |
| None | 509 (70.8) | 18,674 (82.6) |  | 19,183 (82.2) |
| Parent or sibling | 174 (24.2) | 3,560 (15.8) |  | 3,734 (16.0) |
| Parent and sibling | 36 (5.0) | 375 (1.6) |  | 408 (1.8) |
| Prescribed medication, n (%) |  |  |  |  |
| Anti-hypertensive | 114 (15.9) | 2,144 (9.5) | 0.010 | 2,258 (9.7) |
| Steroid | 17 (2.4) | 274 (1.2) | 0.010 | 291 (1.3) |

**Abbreviations:** BMI, body mass index; HbA<sub>1c</sub>, glycated hemoglobin; SD, standard deviation.
Continuous variables are presented as mean ± standard deviation and compared using the independent t-test. Categorical variables are presented as counts (percentages) and compared using the chi-square test.

### 3.2 Correlations among selected omics biomarkers

Figure 1 illustrates the Spearman correlation matrix among the T2D-PRS, selected metabolites, and proteins. Almost no correlation (all |r| < 0.2) was observed between the T2D-PRS and biomarkers from the other omics layers (metabolites and proteins). Correlations between metabolites and proteins were generally weak to moderate (all |r |< 0.5). Within individual omics layers, only a few biomarkers demonstrated strong correlations, defined as |r| ≥ 0.6.

**Figure 1.**
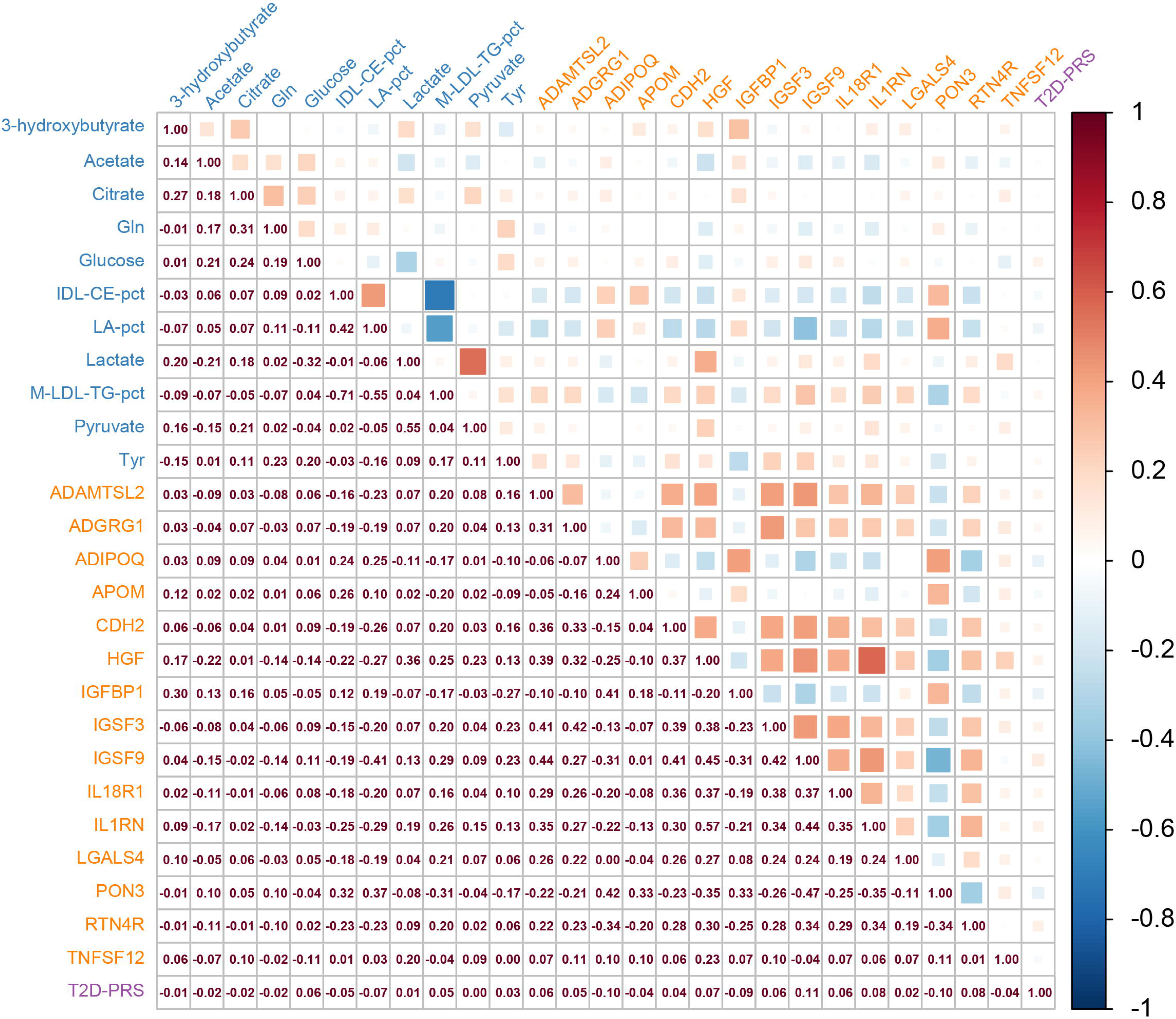
Correlation matrices of selected biomarkers in the validation set (30% of UK Biobank, N=6,998) Colour code for biomarkers: blue, metabolites; orange, proteins; magenta: T2D-PRS. **Abbreviations:** ADAMTSL2, A disintegrin and metalloproteinase with thrombospondin repeats–like 2; ADGRG1, Adhesion G-protein coupled receptor G1; CDH2, Cadherin-2; CDRS, Cambridge Diabetes Risk Score; HGF, Hepatocyte growth factor; IDL-CE-pct, cholesteryl esters to total lipids in IDL percentage; IGFBP1, Insulin-like growth factor-binding protein 1; IGSF, Immunoglobulin superfamily member; IL1RN, Interleukin-1 receptor antagonist; IL18R1, Interleukin-18 receptor 1; LA-pct, linoleic acid to total fatty acids percentage; M- LDL-TG-pct, triglycerides to total lipids in medium LDL percentage; PON3, paraoxonase/lactonase 3; RTN4R, Reticulon-4 receptor; T2D-PRS, polygenic risk score for type 2 diabetes; TNFSF12, TNF-related weak inducer of apoptosis.

### 3.3 Associations of selected omics biomarkers with type 2 diabetes

Figure 2 presents the associations of the selected multi-omics biomarkers with incident type 2 diabetes. Most biomarkers were significantly associated with type 2 diabetes risk, except for five metabolites that did not reach statistical significance. The T2D-PRS was strongly associated with T2D risk (HR per 1 SD: 1.50, 95% CI: 1.32-1.70). Among the metabolomic biomarkers, glucose demonstrated the strongest association with type 2 diabetes (HR per 1 SD: 1.50, 95% CI: 1.34– 1.67) followed by M-LDL-TG-pct (triglycerides to total lipids in medium LDL percentage; HR per 1 SD: 1.46, 95% CI: 1.34–1.60). Among the proteomic biomarkers, IGSF9 (Immunoglobulin superfamily member 9) demonstrated the strongest association with type 2 diabetes (HR [95%CI] per 1 SD): 1.93 [1.72–2.15]).

**Figure 2.**
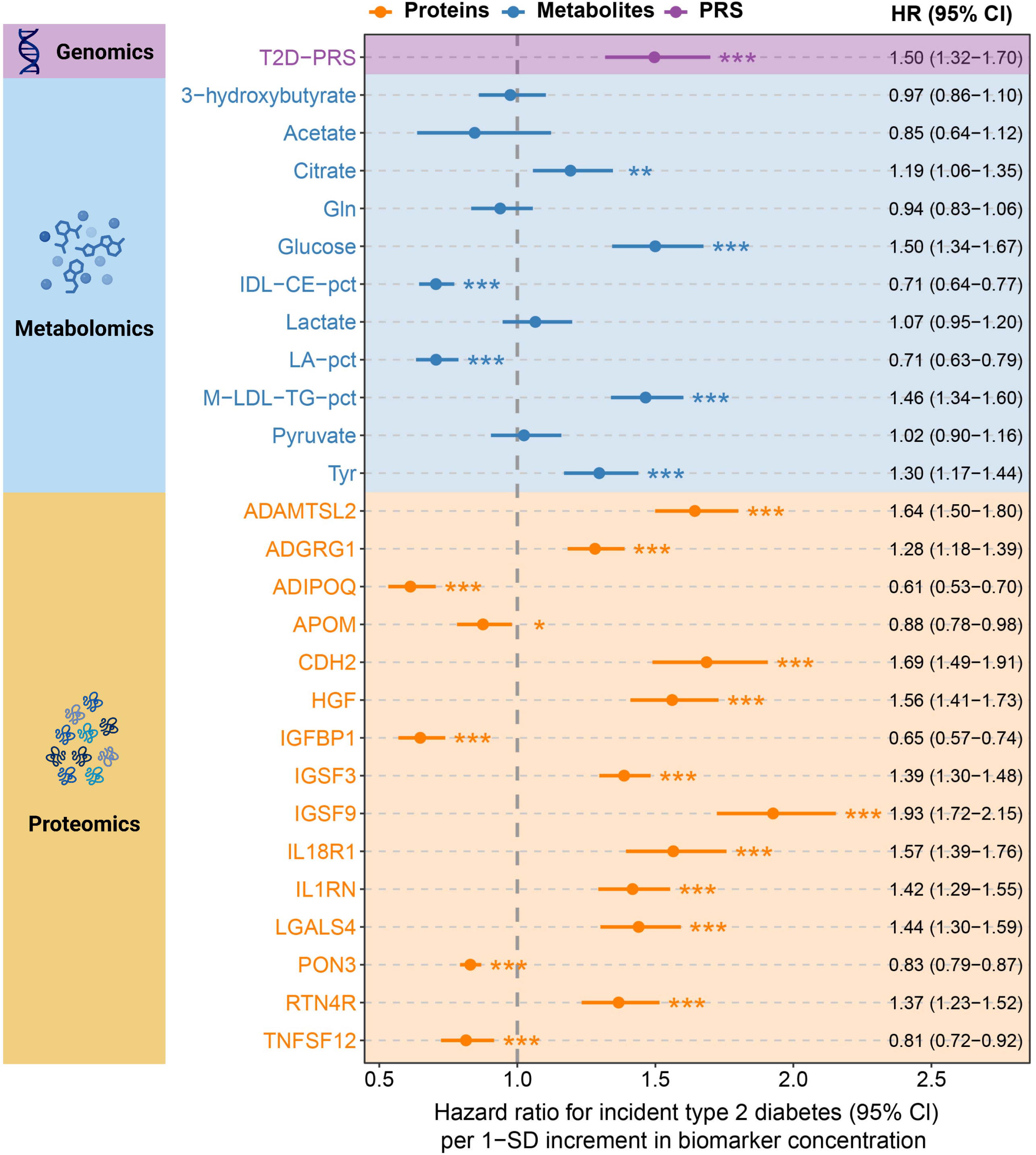
Associations of selected biomarkers with incident type 2 diabetes in the test set (30% of UK Biobank, N = 6,998) Hazard ratios and 95% confidence intervals for one standard deviation increment in biomarker concentration for type 2 diabetes incidence. Analyses were adjusted for all variables included in the clinical CDRS model. Asterisks indicate the level of statistical significance (\**P* < 0.05, \*\**P* < 0.01, \*\*\**P* < 0.001). **Abbreviations:** ADAMTSL2, A disintegrin and metalloproteinase with thrombospondin repeats–like 2; ADGRG1, Adhesion G-protein coupled receptor G1; CDH2, Cadherin-2; CDRS, Cambridge Diabetes Risk Score; HGF, Hepatocyte growth factor; IDL-CE-pct, cholesteryl esters to total lipids in IDL percentage; IGFBP1, Insulin-like growth factor-binding protein 1; IGSF, Immunoglobulin superfamily member; IL1RN, Interleukin-1 receptor antagonist; IL18R1, Interleukin-18 receptor 1; LA-pct, linoleic acid to total fatty acids percentage; M- LDL-TG-pct, triglycerides to total lipids in medium LDL percentage; PON3, paraoxonase/lactonase 3; RTN4R, Reticulon-4 receptor; SD, standard deviation; T2D-PRS, polygenic risk score for type 2 diabetes; TNFSF12, TNF- related weak inducer of apoptosis.

### 3.4 Improvements in Risk Prediction by Multi-Omics Integration

Figure 3 shows the predictive performance of the clinical CDRS model extended by the three multi-omics layers. The C-index of the clinical CDRS model was 0.857. Adding the T2D-PRS (ΔC- index = +0.006; *P*=0.019) or metabolomics data (ΔC-index = +0.013; *P*=0.001) resulted in modest improvements, while integrating proteomics data provided the most substantial improvement (ΔC- index = +0.023; *P*<0.001), underscored by an especially high NRI (30.0%). Thus, in the next step, the proteomics extended clinical CDRS model was chosen as the reference model. Adding either the T2D-PRS or metabolomics to the proteomics-extended model further improved discrimination slightly, but the differences were not statistically significant. However, adding both the T2D-PRS and metabolomics to the proteomics-extended model significantly enhanced discrimination (ΔC- index = +0.006; *P* = 0.033).

**Figure 3.**
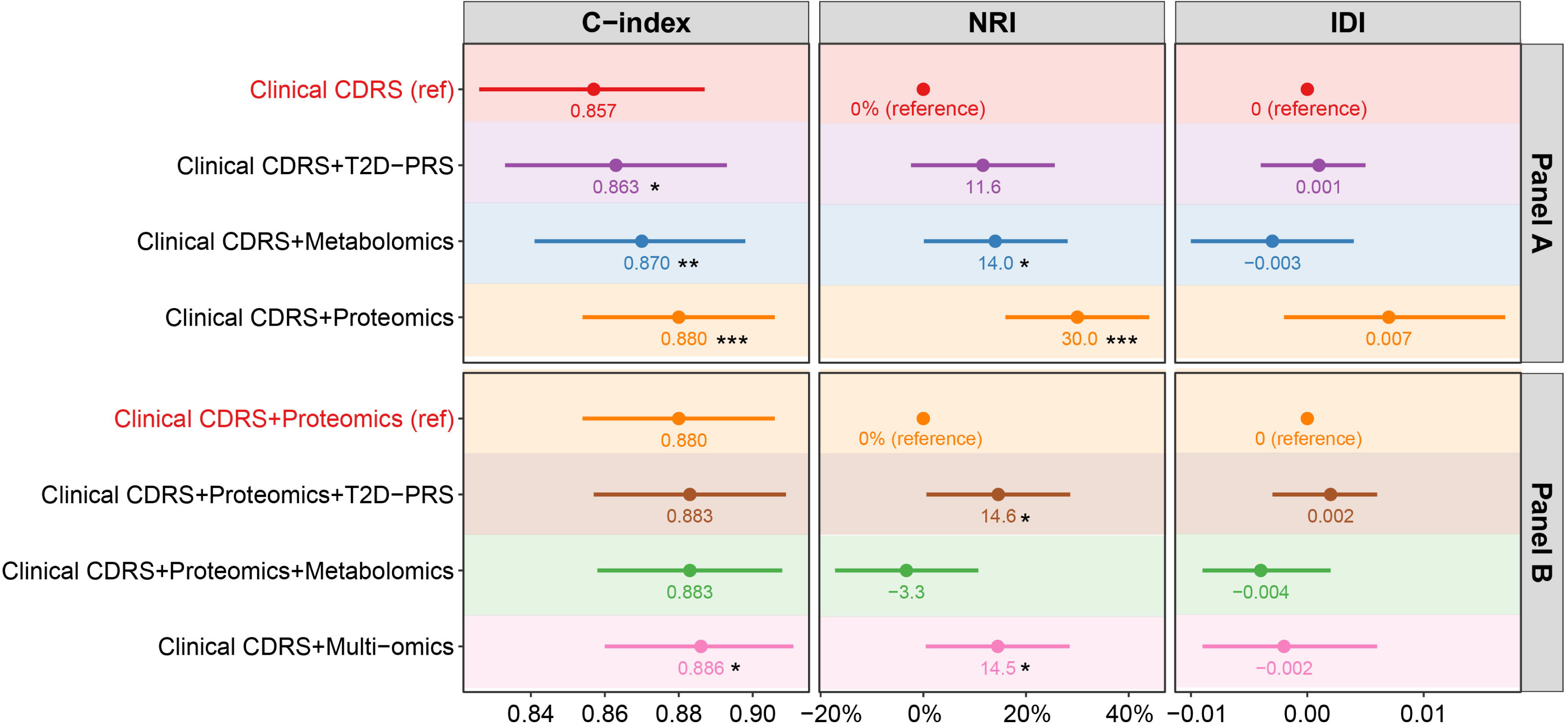
Improvements in 10-year type 2 diabetes risk prediction by integrating multi-omics data in the test set (30% of UK Biobank, N = 6,998) Panel A compares the performance of the clinical CDRS model with its single-omics extensions, including the T2D-PRS, metabolomics, and proteomics. Panel B evaluates the incremental benefit of adding the T2D-PRS or metabolomics to the proteomics-extended model, as well as the full multi-omics model integrating all three layers. Model performance was assessed using C-index, continuous NRI, and IDI. In Panel A, the clinical CDRS served as the reference model. In Panel B, Clinical CDRS + Proteomics served as the reference. Asterisks indicate the statistical significance of performance differences compared with the respective reference model (\**P* < 0.05, \*\**P* < 0.01, \*\*\**P* < 0.001). The multi-omics model refers to the integration of 15 selected proteins, 11 metabolites, and a T2D-PRS. **Abbreviations:** CDRS, Cambridge Diabetes Risk Score; T2D-PRS, polygenic risk score for type 2 diabetes; NRI, net reclassification index; IDI, integrated discrimination improvement.

The ß-coefficients of all variables of the main models (CDRS + Proteomics and CDRS + multi-omics) are detailed in **Supplemental Table S2**. **Supplemental Figure S2** presents the calibration curves for these models, indicating good agreement between predicted and observed event rates across all models.

Figure 4 displays the incremental C-index improvements of each selected biomarker when added individually to the clinical CDRS model. Proteomic biomarkers contributed most strongly to model performance, with 7 of the top 10 contributing biomarkers being proteins, followed by 2 metabolites and the T2D-PRS. Among the proteins, IGSF9 provided the greatest improvement in predictive performance, with an increase in C-index of 0.0156. Among metabolites, M-LDL-TG-pct had the highest predictive contribution, increasing the C-index by 0.0107. The T2D-PRS also showed a significant contribution, increasing the C-index by 0.0060.

**Figure 4.**
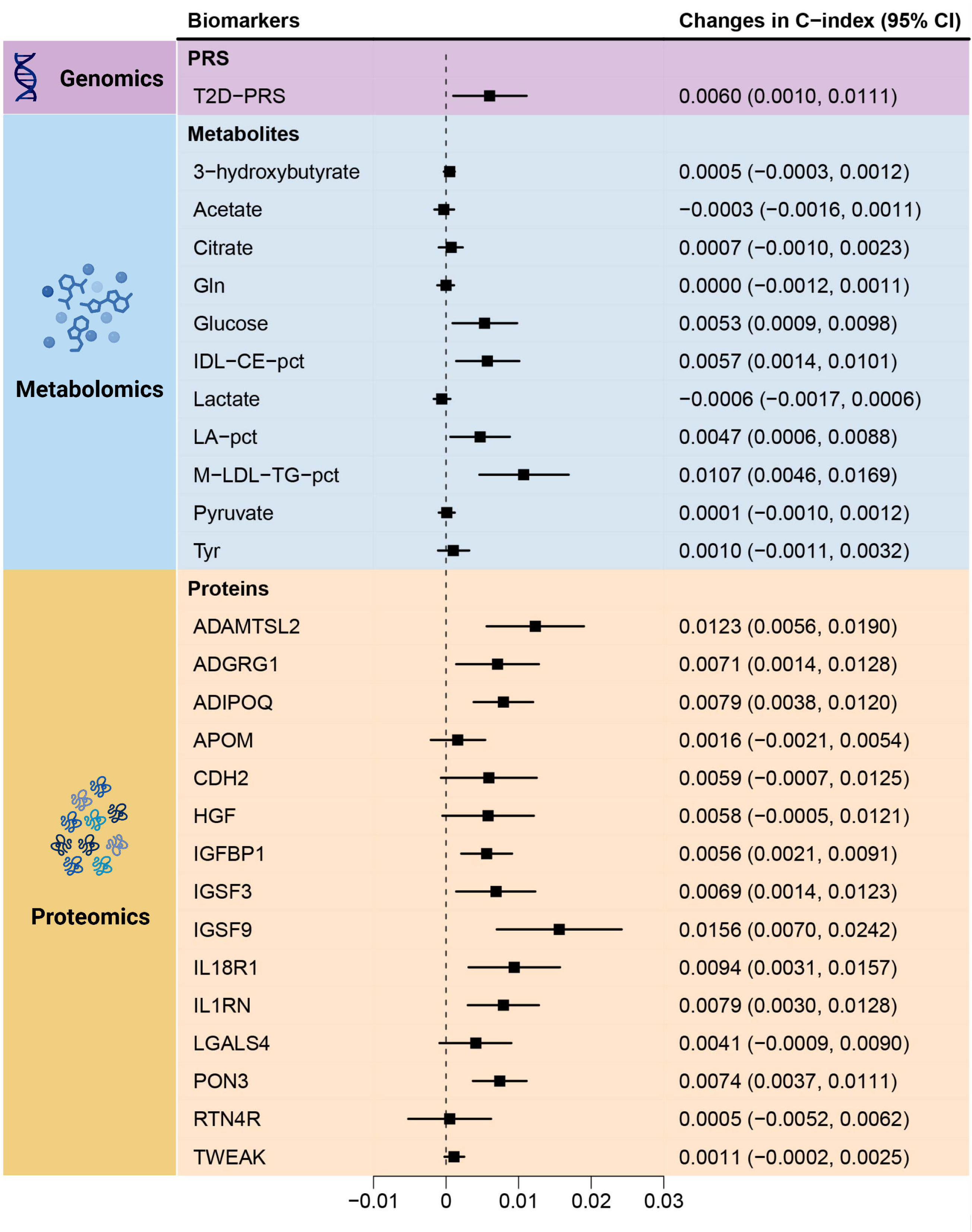
Incremental contributions of individual biomarkers to risk prediction. Forest plot displaying the incremental changes in the C-index when each biomarker is added to the clinical CDRS model. **Abbreviations:** ADAMTSL2, A disintegrin and metalloproteinase with thrombospondin repeats–like 2; ADGRG1, Adhesion G-protein coupled receptor G1; CDH2, Cadherin-2; CDRS, Cambridge Diabetes Risk Score; HGF, Hepatocyte growth factor; IDL-CE-pct, cholesteryl esters to total lipids in IDL percentage; IGFBP1, Insulin-like growth factor-binding protein 1; IGSF, Immunoglobulin superfamily member; IL1RN, Interleukin-1 receptor antagonist; IL18R1, Interleukin-18 receptor 1; LA-pct, linoleic acid to total fatty acids percentage; M- LDL-TG-pct, triglycerides to total lipids in medium LDL percentage; PON3, paraoxonase/lactonase 3; RTN4R, Reticulon-4 receptor; T2D-PRS, polygenic risk score for type 2 diabetes; TNFSF12, TNF-related weak inducer of apoptosis.

In subgroup analyses stratified by prediabetes status, the same patterns emerged in the subjects without prediabetes as in the total population (**Supplemental Figure S3**). Of the three omics layers, proteomics improved the predictive ability of the clinical CDRS most and the full multi-omics model significantly improved this model further. In individuals with prediabetes, adding of proteomics to the considerably increased the C-index from 0.766 (clinical CDRS) to 0.785 but the C-index increases narrowly missed statistical significance (p=0.077). Adding metabolomics data or the T2D-PRS to the proteomics extended clinical CDRS did not further improve prediction among people with prediabetes.

## 4. Discussion

This study investigated whether integrating multi-omics data into a clinical risk model could improve 10-year prediction of type 2 diabetes in a large, population-based cohort of 23,325 adults without previously diagnosed diabetes. We combined a T2D-PRS, 11 metabolites, and 15 proteins with the clinical CDRS and observed significant gains in predictive performance in terms of model discrimination and reclassification. While the full multi-omics model achieved the highest discrimination, its incremental benefit over the proteomics-extended model was modest.

To our knowledge, this is the first large-scale study to systematically evaluate whether integrating genomics, metabolomics, and proteomics improves type 2 diabetes risk prediction beyond single-omics extensions and established clinical models. Previous studies have primarily examined the predictive value of individual omics layers [13, 38–40]. T2D-PRSs have been investigated as early predictors of type 2 diabetes, but their incremental value beyond clinical models has generally been limited [14, 41]. Metabolomics and proteomics have revealed biological pathways relevant to insulin resistance and metabolic dysfunction, and several studies have demonstrated considerable abilities in diabetes risk prediction for these omics data [13, 16, 33, 42–46]. However, these studies focused on single omics layers and did not assess their joint predictive value. In the only prior multi-omics study, conducted in the EPIC-Norfolk cohort (N = 1,105; 375 incident cases), Zanini et al. reported that integration of top features from genomics, proteomics, and metabolomics significantly improved type 2 diabetes prediction compared to the clinical CDRS model (C-index: 0.82 to 0.87; *P* = 0.045), whereas single-omics models showed no significant benefit [18]. Our findings confirm and extend these results in a substantially larger cohort (N = 23,325; 719 incident cases), demonstrating that multi-omics integration significantly enhanced risk prediction (C-index: 0.857 to 0.886). The larger sample size of our cohort allowed more in-depth analyses and elucidated that each omics layer alone significantly improved the predictive abilities of the clinical CDRS, and that proteomics data contributed the largest C-index increase.

Beyond improving overall prediction, multi-omics integration may help identifying individuals who fall below current clinical thresholds for intervention. Existing guidelines recommend preventive action for individuals classified as having prediabetes, defined as HbA_1c_ levels ≥5.7% (≥39 mmol/mol) according to ADA criteria [24]. However, this threshold may fail to identify all individuals at elevated risk for type 2 diabetes and its complications. Our results demonstrate that especially among participants without prediabetes (HbA_1c_ <5.7% [<39 mmol/mol]), the multi-omics model significantly improved risk prediction compared to the clinical CDRS. Thus, multi-omics data may allow early identification of high-risk individuals before prediabetes is clinically detectable. This may be explained by the lower discrimination of the clinical model in individuals without prediabetes, allowing greater room for improvement by biomarkers. In contrast, in those with prediabetes, HbA_1c_ and glucose levels are already strongly predictive, which may limit the added value of omics data.

Recent studies in translational omics have similarly emphasized the need for simplified, interpretable biomarker sets to enable real-world application [47, 48]. While the full multi-omics model achieved the highest overall performance in the total population as well as in people with or without prediabetes, adding proteomics alone achieved almost as good C-index increases in all populations. Thus, the proteomics-extended clinical CDRS model may represent a pragmatic balance between accuracy and feasibility for clinical translation considering the high costs and operational complexity of measuring multi-omics data in clinical settings. In fact, the plasma concentration measurements of only 15 proteins would be needed for the suggested model in this study. The OLINK Focus platform enables simultaneous measurement of up to 21 selected proteins from the broad OLINK Explore library with around 3,000 proteins (https://olink.com/products/olink-focus). A developed 15-protein-panel could offer a cost-effective and scalable solution for implementation in clinical workflows.

This study has several strengths. This is the largest population-based study to evaluate the integration of proteomics, metabolomics, and a T2D-PRS into an established clinical model for type 2 diabetes risk prediction. The large samples size allowed testing each omics layer separately and to conduct a sub-group analysis by prediabetes status. Nonetheless, several limitations should be considered. First, the metabolomics data were derived from a targeted nuclear magnetic resonance platform limited to 250 metabolites, which may not capture the full metabolic complexity relevant to type 2 diabetes. Second, our analyses cannot be generalized to other populations than those with European ancestry aged 37-73 years. Third, external validation in independent cohorts is essential before adoption in routine care. Fourth, although we provide regression coefficients for the selected biomarkers, these may require recalibration when applied to other populations or measurement platforms.

## 5. Conclusions

This study demonstrates for the first time in a large population-based cohort that integrating proteomics, metabolomics, and a T2D-PRS into an established clinical model significantly improves 10-year type 2 diabetes risk prediction beyond single-omics approaches. While the full multi-omics model yielded the highest overall accuracy, the proteomics-extended model alone achieved almost as good model improvements and may represent a more pragmatic option for clinical implementation. These findings highlight the potential of proteomics-based risk models to improve population-level diabetes risk stratification.

## Data Availability

The data that support the findings of this study are available from UK Biobank (https://www.ukbiobank.ac.uk/) but restrictions apply to the availability of these data, which were used under license for the current study (Application Number 101633), and so are not publicly available. Data are however available from the authors upon reasonable request and with permission of UK Biobank.

## List of abbreviations

ADA: American Diabetes Association
ATC: Anatomical Therapeutic Chemical
BMI: Body mass index
CDRS: Cambridge Diabetes Risk Score
CI: Confidence interval
GWAS: Genome-wide association study
HR: Hazard ratio
IDI: Integrated discrimination improvement
IDL-CE-pct: cholesteryl esters to total lipids in IDL percentage
IGSF9: Immunoglobulin superfamily member 9
LASSO: Least absolute shrinkage and selection operator
M-LDL-TG-pct: triglycerides to total lipids in medium LDL percentage
NMR: Nuclear magnetic resonance
NRI: Net reclassification index
PRS: Polygenic risk score
T2D-PRS: Type 2 diabetes polygenic risk score
UKB: UK Biobank.

## Acknowledgements

This research has been conducted using the UK Biobank Resource under Application Number 101633. This work uses data provided by patients and collected by the NHS as part of their care and support. We would like to thank all participants of the UK Biobank as well as the staff of the UK Biobank assessment centers for their contributions.

## CRediT authorship contribution statement

**Ruijie Xie:** Writing – original draft, Investigation, Formal analysis, Software, Data curation, Conceptualization. **Christian Herder:** Writing – review & editing. **Ben Schöttker:** Writing – review & editing, Supervision, Investigation, Methodology, Data curation, Conceptualization, Funding acquisition.

---

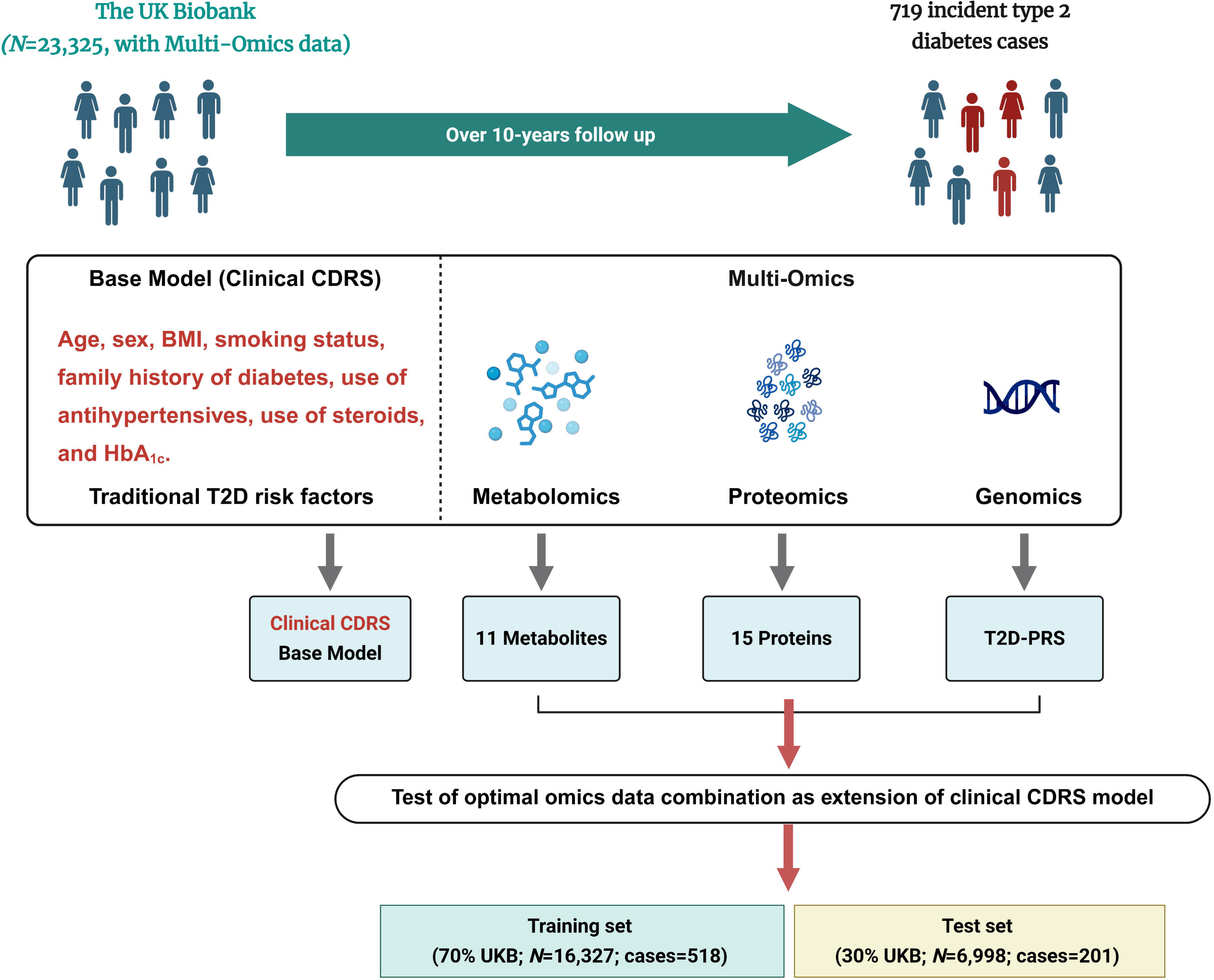

