## Supplemental Materials for "Large-Scale Multi-Omics Enhance Risk Prediction for Type 2 Diabetes"

**Supplemental Table S1.** Selected multi-omic biomarkers and data sources

| **Omics** | **Biomarkers** | **Reference** |
| --- | --- | --- |
| PRS | Standard PRS for type 2 diabetes | The standard T2D-PRS was derived using a genome-wide meta-analysis of 5 external GWAS datasets comprising 1,349,223 individuals and has been previously published [1]. |
| Metabolites | 3-hydroxybutyrate, acetate, citrate, Gln, glucose, IDL-CE-pct, LA-pct, lactate, M-LDL-TG-pct, pyruvate, Tyr | The metabolites were selected using bootstrap-LASSO regression in the UKB (n=60,362). See previous work [2]. |
| Proteins | ADAMTSL2, ADGRG1, ADIPOQ, APOM, CDH2, HGF, IGFBP1, IGSF3, IGSF9, IL18R1, IL1RN, LGALS4, PON3, RTN4R, TNFSF12 | The proteins were selected using bootstrap-LASSO regression in the UKB (n=14,759). See previous work [3]. |

Metabolites were quantified from EDTA plasma using nuclear magnetic resonance spectroscopy, and proteins were measured from EDTA plasma using Olink proximity extension assays.

**Abbreviations:** ADAMTSL2, A disintegrin and metalloproteinase with thrombospondin repeats–like 2; ADGRG1, Adhesion G-protein coupled receptor G1; CDH2, Cadherin-2; CDRS, Cambridge Diabetes Risk Score; HGF, Hepatocyte growth factor; IDL-CE-pct, cholesteryl esters to total lipids in IDL percentage; IGFBP1, Insulin-like growth factor-binding protein 1; IGSF, Immunoglobulin superfamily member; IL1RN, Interleukin-1 receptor antagonist ; IL18R1, Interleukin-18 receptor 1; LA-pct, linoleic acid to total fatty acids percentage; M-LDL-TG-pct, triglycerides to total lipids in medium LDL percentage; PON3, paraoxonase/lactonase 3; PRS, polygenic risk score; RTN4R, Reticulon-4 receptor; T2D, type 2 diabetes; T2D-PRS, polygenic risk score for type 2 diabetes; TNFSF12, TNF-related weak inducer of apoptosis.

**Supplemental** **Table S2.** ß-coefficients of the different extensions of the clinical CDRS model with multi-omics data for 10-year prediction of type 2 diabetes

| **Risk factor (units)** | **ß-coefficient** | |
| --- | --- | --- |
| **Clinical CDRS variables** | **Clinical CDRS + Proteomics** | **Clinical CDRS + Multi-omics** |
| Female | -0.3040 | -0.2743 |
| Prescribed anti-hypertension medication | 0.0334 | 0.0168 |
| Prescribed steroids | 0.4104 | 0.4647 |
| Age (per years) | 0.0077 | 0.0077 |
| Body mass index <25 kg/m^2^ | Ref | Ref |
| Body mass index 25-27.49 kg/m^2^ | -0.0833 | -0.1195 |
| Body mass index 27.5-29.99 kg/m^2^ | 0.2421 | 0.1798 |
| Body mass index ≥ 30 kg/m^2^ | 0.3493 | 0.3178 |
| No first degree relative with diabetes | Ref | Ref |
| Parent or sibling with diabetes | 0.2846 | 0.2655 |
| Parent and sibling with diabetes | 0.5445 | 0.4719 |
| None-smoker | Ref | Ref |
| Ex-smoker | 0.1552 | 0.1504 |
| Current smoker | -0.0466 | -0.0350 |
| HbA_1c_ (per %) | 2.6941 | 2.4053 |
| **Added proteins (per 1 SD)** |  |  |
| ADAMTSL2 | 0.0262 | 0.0360 |
| ADGRG1 | 0.0316 | 0.0297 |
| ADIPOQ | 0.0642 | 0.0905 |
| APOM | -0.0930 | -0.0876 |
| CDH2 | 0.1098 | 0.0817 |
| HGF | 0.1062 | 0.1669 |
| IGFBP1 | -0.1341 | -0.1308 |
| IGSF3 | -0.0043 | -0.0206 |
| IGSF9 | 0.1384 | 0.0878 |
| IL18R1 | 0.1452 | 0.1194 |
| IL1RN | 0.0389 | 0.0532 |
| LGALS4 | 0.1180 | 0.0993 |
| PON3 | -0.1297 | -0.1084 |
| RTN4R | 0.1406 | 0.1595 |
| TNFSF12 | -0.0615 | -0.0672 |
| **Additional metabolites** **(per 1 SD)** |  |  |
| 3-hydroxybutyrate | - | -0.0144 |
| Acetate | - | 0.0186 |
| Citrate | - | 0.0300 |
| Gln | - | 0.0479 |
| Glucose | - | 0.2071 |
| IDL-CE-pct | - | -0.0088 |
| LA-pct | - | -0.0751 |
| Lactate | - | 0.0602 |
| M-LDL-TG-pct | - | 0.0549 |
| Pyruvate | - | -0.1157 |
| Tyr | - | -0.0469 |
| **Additional PRS** **(per 1 SD)** |  |  |
| T2D-PRS | - | 0.2988 |

**Abbreviations:** ADAMTSL2, A disintegrin and metalloproteinase with thrombospondin repeats–like 2; ADGRG1, Adhesion G-protein coupled receptor G1; ADIPOQ, Adiponectin; APOM, Apolipoprotein M; CDH2, Cadherin-2; CDRS, Cambridge Diabetes Risk Score; HGF, Hepatocyte growth factor; IDL-CE-pct, cholesteryl esters to total lipids in IDL percentage; IGFBP1, Insulin-like growth factor-binding protein 1; IGSF, Immunoglobulin superfamily member; IL1RN, Interleukin-1 receptor antagonist; IL18R1, Interleukin-18 receptor 1; LA-pct, linoleic acid to total fatty acids percentage; M-LDL-TG-pct, triglycerides to total lipids in medium LDL percentage; PON3, paraoxonase/lactonase 3; PRS, polygenic risk score; Ref, reference; RTN4R, Reticulon-4 receptor; SD, standard deviation; T2D, type 2 diabetes; TNFSF12, TNF-related weak inducer of apoptosis.

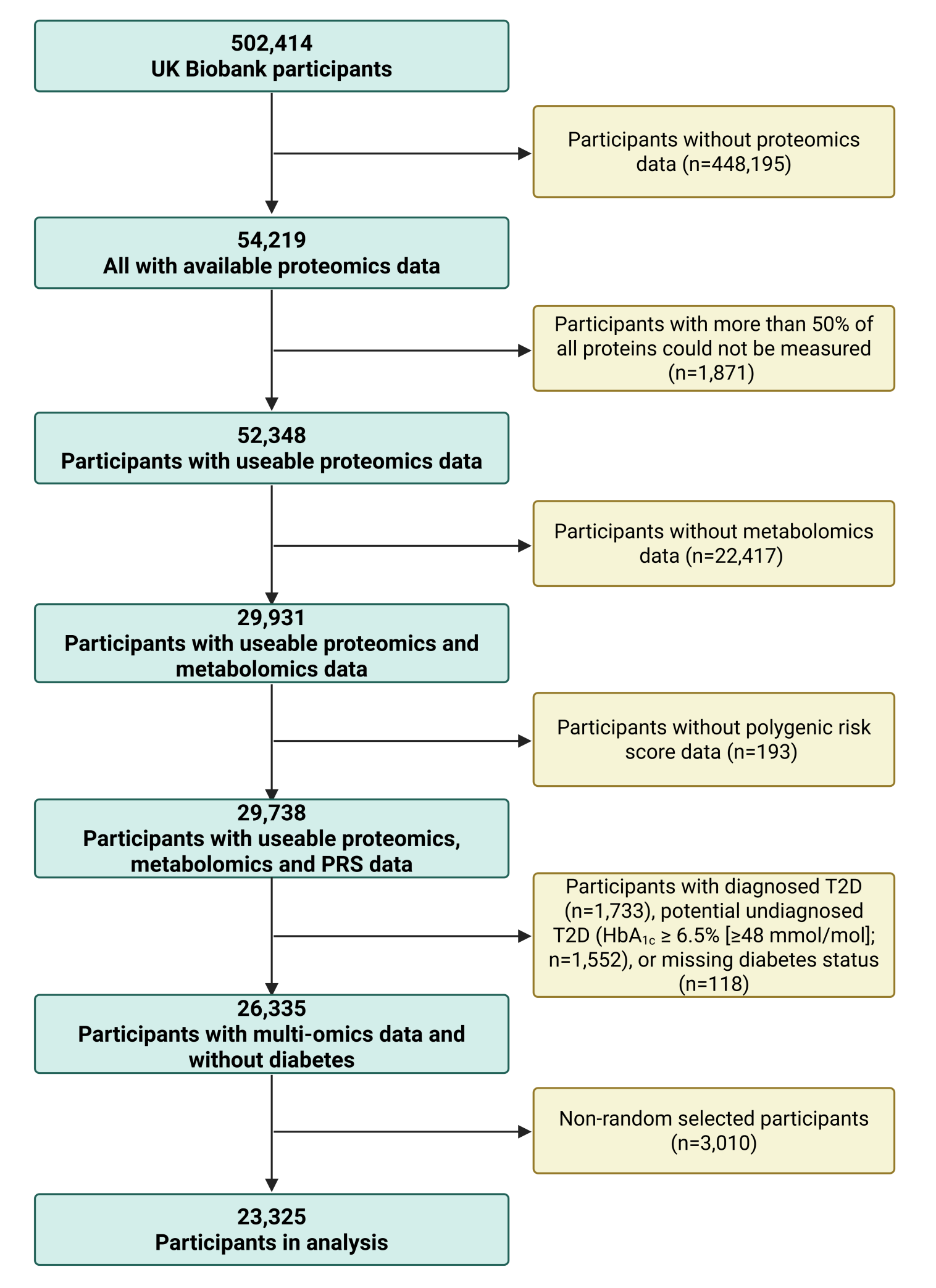

**Supplemental Figure S1.** Flowchart of participant selection

**Abbreviations:** HbA_1c_, glycated hemoglobin; PRS, polygenic risk score; T2D, type 2 diabetes.

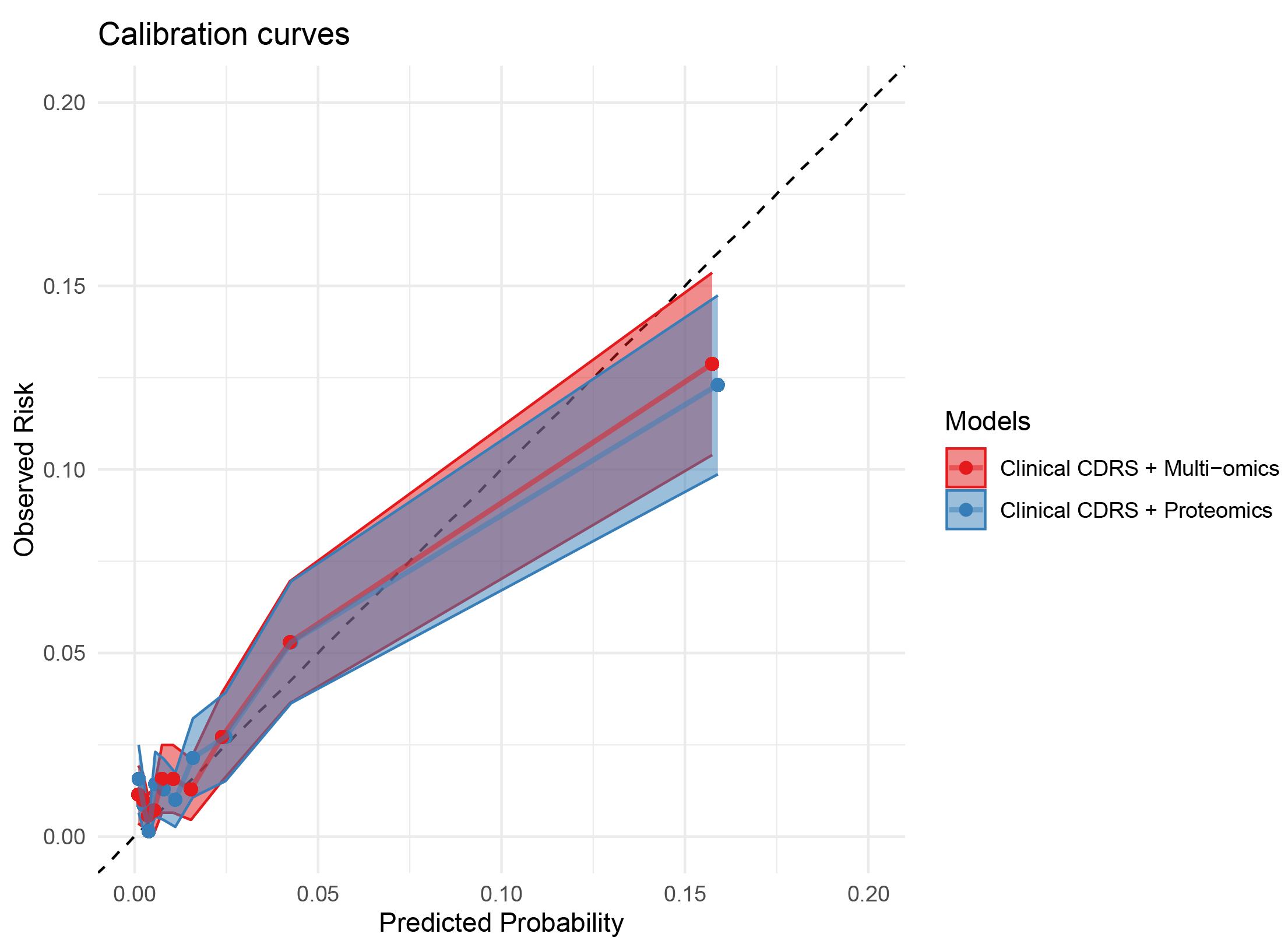

**Supplemental Figure S2.** Calibration curves of the different extensions of the clinical CDRS model with multi-omics data for 10-year type 2 diabetes risk prediction in the test set (30% of UK Biobank, N=6,998)

The multi-omics model refers to the integration of proteomics, metabolomics, and the T2D-PRS.

**Abbreviations:** CDRS, Cambridge Diabetes Risk Score.

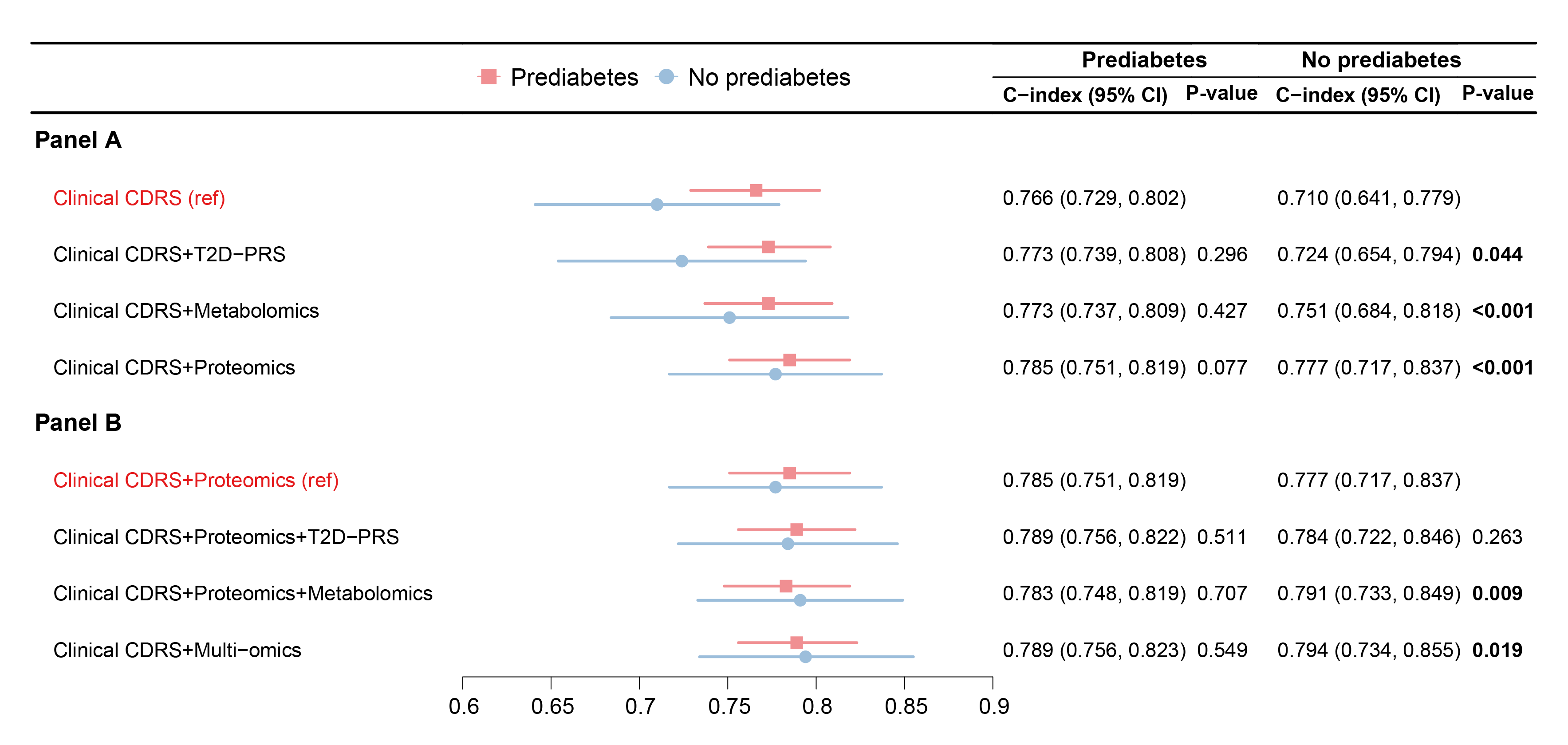

### **Supplemental Figure S3.** Subgroup analysis of predictive performance by prediabetes status (30% of UK Biobank, N=6,998)

The number of individuals and cases was 1,009 (135 cases, 874 non-cases) in the group with prediabetes and 5,989 (66 cases, 5,923 non-cases) in the group without prediabetes.

Panels A and B show the C-index and 95% confidence intervals for the clinical CDRS model and its multi-omics extensions in two subgroups: individuals without prediabetes (HbA_1c_ < 5.7% [< 39 mmol/mol]; Panel A) and with prediabetes (HbA_1c_ ≥ 5.7% [≥ 39 mmol/mol]; Panel B), based on ADA criteria. In Panel A, the clinical CDRS was used as the reference model; in Panel B, Clinical CDRS + Proteomics served as the reference.

All P values represent comparisons of the C-index with the respective reference model. Values in bold indicate P < 0.05.

The multi-omics model refers to the integration of proteomics, metabolomics, and the T2D-PRS.

**Abbreviations:** CDRS, Cambridge Diabetes Risk Score; T2D-PRS, polygenic risk score for type 2 diabetes.

**References to Supplemental Materials**

1.	Thompson DJ, Wells D, Selzam S, Peneva I, Moore R, Sharp K, Tarran WA, Beard EJ, Riveros-Mckay F, Giner-Delgado C *et al*: **A systematic evaluation of the performance and properties of the UK Biobank Polygenic Risk Score (PRS) Release**. *PLoS One* 2024, **19**(9):e0307270.

2.	Xie R, Herder C, Sha S, Peng L, Brenner H, Schoettker B: **Novel type 2 diabetes prediction score based on traditional risk factors and circulating metabolites: Model derivation and validation in two large cohort studies**. *eClinicalMedicine* 2025, **79**.

3.	Xie R, Vlaski T, Trares K, Herder C, Holleczek B, Brenner H, Schöttker B: **Large-Scale Proteomics Improve Risk Prediction for Type 2 Diabetes**. *Diabetes Care* 2025.
